# A Cross-sectional Study of Clinical COVID-19 Myocarditis: Differences in Biomarkers in Fulminant and Non-fulminant Cases

**DOI:** 10.1101/2021.06.06.21258423

**Authors:** Christopher Wong, Amtul Mansoor, Thomas McGinn

**Affiliations:** Department of Medicine, North Shore University Hospital & Long Island Jewish Medical Center, Manhasset, NY, USA; Donald and Barbara Zucker School of Medicine at Hofstra/Northwell, Hempstead, NY, USA; CommonSpirit Health, Chicago, IL, USA

**Keywords:** COVID-19, myocarditis

## Abstract

**Background:** COVID-19 myocarditis is becoming increasingly appreciated as a complication of COVID-19. There are significant hurdles to formal diagnosis with endomyocardial biopsy or cardiac MRI whether by resource limitations, patient instability, or isolation precautions. Therefore, further exploratory analysis is needed to clinically define the characteristics and spectrum of severity of COVID-19 myocarditis.

**Objectives:** The aim of this study was to describe the clinical course, echocardiographic, and laboratory testing across suspected fulminant and non-fulminant clinically defined COVID-19 myocarditis.

**Methods:** In a cross-sectional observational study of 19 patients with clinically defined COVID-19 myocarditis, we report presenting symptoms, clinical course, laboratory findings, and echocardiographic results stratified by non-fulminant and fulminant myocarditis. Student t-test and univariate logistic regression are used to compare laboratory findings across fulminant and non-fulminant cases.

**Findings:** Among 19 patients, there was no prior history of coronary artery disease, atrial fibrillation, or heart failure; 21.1% of patients died; and 78.9% of cases required supplemental oxygen. A significantly higher geometric mean D-dimer and ferritin were observed in patients with fulminant compared to non-fulminant suspected myocarditis. 26.3% of cases had pericardial effusions. 10 out of the 16 with available echocardiographic data had normal left ventricular systolic function.

**Conclusions:** In this cross-sectional analysis, we provide a practical clinical depiction of patients with clinical COVID-19 myocarditis across fulminant and non-fulminant cases. Statistically significant elevations in inflammatory markers in fulminant versus non-fulminant cases generate hypothesis regarding the role of systemic inflammation in driving severity of COVID-19 myocarditis.

## INTRODUCTION

Coronavirus disease 2019 (COVID-19) caused by the coronavirus 2 (SARS-CoV-2) predominantly affects the respiratory system but may also result in myocarditis.^1^ SARS-CoV-2 binds to the angiotensin converting enzyme-2 receptor expressed on the surface of alveolar and cardiac cells, which may account for the direct cardiac involvement in COVID-19.^2^ Myocarditis is an inflammatory disease caused by infectious and non-infectious etiologies.^3^ Fulminant myocarditis is the most severe type of myocarditis and is predominantly caused by viral infections. It is characterized by a sudden and severe inflammation of myocardium resulting in cardiogenic shock, ventricular tachyarrhythmias or bradyarrythmias.^3^

Clinical presentation of patients with COVID-19 myocarditis varies greatly, but signs and symptoms include fatigue, dyspnea, chest pain, acute-onset heart failure, and cardiogenic shock.^4^ Previous studies have reported elevated levels of inflammatory markers, such as C-reactive protein (CRP), erythrocyte sedimentation rate (ESR), procalcitonin, and lactate. Patients also had elevated levels of troponins and pro-BNP levels.^5^ A variety of electrocardiogram (ECG) abnormalities can be seen in patients with myocarditis ranging from ST elevation and PR depression to new-onset bundle branch block, QT prolongation, brady- or tachy-arrhythmias.^6^

In patients with clinical suspicion for myocarditis, American Heart Association (AHA) recommends further testing with echocardiogram or cardiovascular magnetic resonance (CMR).^7^ On echocardiogram, patients with myocarditis have increased wall thickness, chamber dilation, and pericardial effusion, and ventricular systolic dysfunction.^7^ CMR can provide excellent insights into tissue-level pathologies but it’s utility is limited by availability and poor image quality with tachycardia, and requirements for deep cleaning after each use in the midst of epidemics and pandemics. The gold standard for diagnosis of myocarditis in endomycardial biopsy (EMB). EMB biopsies are limited due to the risk of contagious spread, the expertise required and false negative rate.^4^ They have only been recommended for confirmation of myocarditis in severe cases.^8^

Further work is needed to better characterize suspected COVID-19 myocarditis as a clinical entity, because instability, infection control, and resource limitations may prohibit timely, definitive diagnosis by CMR or EMB especially in low-resource settings where the spread of COVID-19 continues. The differential for multiorgan failure and hypotension is overshadowed by ARDS and septic shock; bilateral pulmonary infiltrates are more likely to be attributed to primary pulmonary pathology rather than congestive heart failure; and decreased left ventricular function and troponin elevation may be attributed to demand mediated ischemia or sepsis induced cardiomyopathy. The clinical significance of COVID-19 myocarditis in the majority of cases could be easily overlooked.

However, cardiac inflammation with elevated CRP and troponins compared to controls have been noted in patients recovered from COVID-19 and multiple case reports of COVID-19 myocarditis have been previously reported.^9,10^ 17% of patients with COVID-19 had troponin elevations above the 99th percentile while only 2.9% of patients hospitalized with influenza had troponin elevations.^11,12^ In summary, there is good reason to believe that COVID-19 is uniquely cardiotropic yet the methods for rigorous diagnosis are typically unavailable. COVID-19 myocarditis may be an overlooked clinical entity with potentially significant impact.

In order to further characterize clinical COVID-19 myocarditis as an entity and delineate the spectrum of disease across non-fulminant and fulminant COVID-19 myocarditis, we present a cross-sectional observational study of 19 patients with COVID-19 who were clinically diagnosed with myocarditis based on presenting symptoms, lab values, and echocardiogram.

## METHODS

The charts of 41 patients admitted to a hospital in the Northwell Health system in the state of New York between March 21, 2020 and May 18, 2020 were reviewed. 6 of these 41 patients were referred directly to the researchers’ attention. The remaining 35 were found by querying the system-wide Northwell electronic health record for admitted patients with retropharyngeal swabs positive for SARS-CoV-2 by polymerase chain reaction and ICD-10 codes with the word “myocarditis” in their description (I51.4, I40.0, I41, B33.22, I40.9). This was a convenience sample without a predetermined sample size.

Based on guidelines from European Society of Cardiology, clinical COVID-19 myocarditis was defined as meeting at least one out of four signs of clinical presentation in addition to one diagnostic criterion and the absence of other clinical conditions that could explain the clinical findings. The clinical presentations were acute chest pain, new onset or worsening dyspnea and/or fatigue, palpitations, and unexplained cardiogenic shock. The diagnostic criteria included: ECG changes with 1^st^ to 3^rd^ degree AV block, ST/T wave changes (STE or TWI), sinus arrest, ventricular tachycardia or fibrillation, atrial fibrillation, intraventricular conduction delay (widened QRS complex), supraventricular tachycardia; elevated troponins; and functional and structural abnormalities on echocardiogram.^13^ Patients were excluded if acute coronary syndrome was deemed more likely than myocarditis. 19 of the 41 patients reviewed met these criteria for suspected myocarditis. Cases were categorized as fulminant or non-fulminant where fulminant cases were defined as myocarditis with new onset heart failure requiring ionotropic or mechanical circulatory support.

We report the baseline characteristics, outcomes, laboratory findings, and echocardiographic findings of these 19 patients. We report age, sex, body mass index, presence of shortness of breath or chest pain on admission, and supplemental oxygen requirements. Presence of shortness of breath and chest pain were ascertained by chart review of patient’s documented symptoms. Supplemental oxygen was defined as either room air, nasal canula, nonrebreather, or intubation. High-flow nasal canula and bilevel positive airway protection were being avoided during the analyzed time period due to infection control policies in response to the pandemic. We also report any recorded history of hypertension, type 2 diabetes, atrial fibrillation, coronary artery disease, heart failure, and chronic kidney. The mean and standard deviation are reported for continuous variables while the counts and percentages are reported for categorical variables. Findings are stratified by fulminant and non-fulminant cases.

We report laboratory data at the time of diagnosis. When available, the highest values for troponin, CRP, lactate, D-dimer, ferritin, and procalcitonin within the first 48 hours of clinical COVID-19 myocarditis were recorded. We report troponin as the fold increase of the upper limit of normal because some facilities used troponin I and others troponin T. The upper limit of troponin T was defined as 51 ng/L and the upper limit of troponin I was defined as 15 ng/L. The arithmetic mean and standard deviation are reported for all lab values. We report the geometric mean of the CRP, lactate, D-dimer, ferritin, and procalcitonin. All measurements are stratified by fulminant and non-fulminant myocarditis. We chose to estimate the central tendency of the inflammatory measurements by the geometric mean because these markers represent a multiplicative biologic process of positive feedback loops thought to drive the highly inflammatory state in severe COVID-19.^14^ Geometric mean was not reported for troponins because of values equal to 0. Missing data are reported.

As a means of quantifying descriptive statistical difference, two-sided student t-test was used to evaluate for statistically significant differences across all arithmetic means across fulminant and non-fulminant cases. Univariate regression of the log-transformed measurement over fulminant and non-fulminant status were used to evaluate the significance of the difference in geometric mean. P-values are reported.

We reviewed echocardiographic data and report ejection fraction, presence of left ventricular systolic dysfunction, left ventricular diastolic dysfunction, and the presence of pericardial effusion. Ejection fraction was reported as an ordinal variable (15-40%, 40-50%, and greater than 50%). Left ventricular systolic dysfunction were reported either based on formal echocardiography or critical care point of care ultrasound. Diastolic dysfunction and pericardial effusions were only reported based off formal echocardiography. Missing data are reported.

The study received expedited IRB approval and was deemed within the scope of IRB-approved data registry for patients with COVID-19 at Northwell Health.

Analysis was done with R and the uwintrostats and TableOne packages.

## RESULTS

Analysis of demographic information of 19 patients with clinical COVID-19 myocarditis showed mean age was 58.42 years, 68.4% of patients were male, and mean body mass index was 30.13. There was a lower proportion of males (62.5% v 80%) and a higher average BMI (31.91 v 28.70) in the non-fulminant group. 94.7% patients endorsed shortness of breath on presentation while only 26.3% endorsed chest pain. Chest pain was more common in non-fulminant cases (40% v 11.1%) (Table 1).

**Table 1:** Demographic and Past Medicals History of 19 Patients with Clinical COVID-19 Myocarditis

|  | Non-fulminant | Fulminant | Overall |
| --- | --- | --- | --- |
| n | 10 | 9 | 19 |
| Age (mean (SD)) | 61.90 (15.84) | 54.56 (20.49) | 58.42 (18.06) |
| Male (n (%)) | 5 ( 50.0) | 8 ( 88.9) | 13 ( 68.4) |
| Body Mass Index (kg/m<sup>2</sup>) (mean (SD))<sup>1</sup> | 31.50 (6.48) | 28.76 (5.66) | 30.13 (6.07) |
| Presented with Shortness of Breath (n (%)) | 10 (100.0) | 8 ( 88.9) | 18 ( 94.7) |
| Presented with Chest Pain (n (%)) | 4 ( 40.0) | 1 ( 11.1) | 5 ( 26.3) |
| History of Hypertension (n (%)) | 7 ( 70.0) | 3 ( 33.3) | 10 ( 52.6) |
| History of Type 2 Diabetes (n (%)) | 5 ( 50.0) | 2 ( 22.2) | 7 ( 36.8) |
| History of Atrial Fibrillation (n (%)) | 0 (0) | 0 (0) | 0 (0) |
| History of Coronary Artery Disease (n (%)) | 0 (0) | 0 (0) | 0 (0) |
| History of Heart Failure | 0 (0) | 0 (0) | 0 (0) |
| History of Chronic Kidney Disease (n (%)) | 3 ( 30.0) | 1 ( 11.1) | 4 ( 21.1) |
| Supplemental oxygen (n (%)) |  |  |  |
| Room air | 3 (30.0) | 1 (11.1) | 4 (21.1) |
| Nasal Canula | 3 (30.0) | 2 (22.2) | 5 (26.3) |
| Nonrebreather | 2 (20.0) | 0 ( 0.0) | 2 (10.5) |
| Intubation | 2 (20.0) | 6 (66.7) | 8 (42.1) |
| Length of stay in days (mean (SD)) | 10.66 (8.74) | 20.17 (13.60) | 15.17 (12.01) |
| Death (n (%)) | 2 ( 20.0) | 2 ( 22.2) | 4 ( 21.1) |

In terms of cardiovascular risk factors, 52.6% of patients had hypertension and 36.8% had type 2 diabetes. Hypertension and type 2 diabetes tended to be more prevalent in patients with non-fulminant myocarditis (70% v 33.3%; 50% v 22.2%). There was no documented history of atrial fibrillation, coronary artery disease, or prior heart failure. Overall, 21.1% of patients had chronic kidney disease. Regarding their clinical course, a lower proportion of patients with non-fulminant myocarditis were intubated (20% v 66.7%). Overall, 21.1% of patients died during hospitalization with grossly similar mortality between non-fulminant and fulminant cases (20% v 22.2%) (Table 1).

In exploratory analysis of laboratory findings at the time of suspecting COVID-19 myocarditis, mean fold elevation in troponin above normal limit (100.84), CRP (80 mg/L), lactate (2.79 mmol/L), D-dimer (3775 ng/mL), ferritin (8355 ng/mL), and procalcitonin (23 ng/mL) were all markedly elevated. With one patient with missing data, mean fold-increase of troponins were non-significantly higher in non-fulminant cases than fulminant cases (101 v 71, p = .303). With 2 cases with missing data, mean and geometric mean of lactates were similar between non-fulminant and fulminant cases (2.76 mmol/L v 2.82 mmol/L, p = .72; 2.64 mmol/L v 2.14 mmol/L, p =.83). With 4 cases that had missing data, fulminant cases had non-significantly higher mean D-dimer and significantly higher geometric mean D-dimer (2609 ng/mL v 4796 ng/mL, p = .253; 1199 ng/mL v 4101 ng/mL, p = .028). With 4 cases that had missing data, fulminant cases had non-significantly higher mean ferritin and significantly higher geometric mean ferritin (1702 ng/mL v 14176 ng/mL, p = .089; 1468 ng/mL v 5660 ng/mL, p = .046) (Table 2).

**Table 2:** Laboratory findings at time of clinical COVID-19 myocarditis

|  | Mean (sd, number missing) |  |  |  | Geometric Mean (number missing) |  |  |  |
| --- | --- | --- | --- | --- | --- | --- | --- | --- |
|  | Non-fulminant | Fulminant | Overall | p-value | Non-fulminant | Fulminant | Overall | p-value |
| n | 9 | 10 | 19 |  | 9 | 10 | 19 |  |
| Fold increase in troponin above normal limit | 71 (91.61, 1) | 131 (144.14, 0) | 101 (121.23, 1) | 0.303 | NA | NA | NA |  |
| C-reactive Protein (mg/L) | 88 (96.41, 1) | 71 (100.20, 2) | 79 (95.43, 3) | 0.725 | 39 (1) | 45 (2) | 42 (3) | 0.83 |
| Lactate (mmol/L) | 2.76 (0.85, 2) | 2.82 (2.64, 0) | 2.79 (1.95, 2) | 0.952 | 2.64 (2) | 2.14 (0) | 2.36 (2) | 0.45 |
| D-dimer (ng/mL) | 2609 (4533.78, 3) | 4796 (2367.48, 1) | 3775 (3589.89, 4) | 0.253 | 1199 (3) | 4101 (1) | 2311 (4) | 0.028 |
| Ferritin (ng/mL) | 1702 (910.39, 3) | 14176 (17836.06, 1) | 8355 (14174.19, 4) | 0.089 | 1468 (3) | 5660 (1) | 3015 (4) | 0.046 |
| Procalcitonin (ng/mL) | 1.76 (1.40, 4) | 37.74 (72.92, 0) | 23.35 (58.07, 4) | 0.254 | 1.08 (4) | 6.74 (0) | 3.24 (4) | 0.067 |

Descriptive statistics of echocardiographic data shows that all cases of fulminant myocarditis have decreased systolic dysfunction as expected, but ejection fractions less than 40% were not observed (Table 3). Of the 10 non-fulminant cases, 8 had data on ejection fractions. 4 of these patients had preserved ejection fractions greater than 50% while 2 had ejection fractions between 15% and 40%. Of the 9 cases of suspected fulminant myocarditis, 5 had data on ejection fraction all of which had ejection fractions between 40 and 50%. Only 1 case demonstrated diastolic dysfunction and 5 cases were observed to have pericardial effusions out of 16 cases that were evaluated with echocardiography.

**Table 3:** Echocardiographic findings of Clinical COVID-19 Myocarditis

| Table 3: Echocardiographic findings of SARS-COV-2 Myocarditis |  |  |  |
| --- | --- | --- | --- |
|  | Non-fulminant | Fulminant | Overall |
| n | 10 | 9 | 19 |
| Ejection Fraction (n (%)) |  |  |  |
| >50 | 5 (50.0) | 0 ( 0.0) | 5 (26.3) |
| 40-50 | 1 (10.0) | 5 (55.6) | 6 (31.6) |
| 15-40 | 2 (20.0) | 0 ( 0.0) | 2 (10.5) |
| Missing data | 2 (20.0) | 4 (44.4) | 6 (31.6) |
| Left Ventricular Systolic Dysfunction (n (%)) |  |  |  |
| Left ventricular systolic dysfunction present | 2 (20.0) | 8 (88.9) | 10 (52.6) |
| No left ventricular systolic dysfunction | 6 (60.0) | 0 ( 0.0) | 6 (31.6) |
| Missing data | 2 (20.0) | 1 (11.1) | 3 (15.8) |
| Left Ventricular Diastolic Dysfunction (n (%)) |  |  |  |
| Left ventricular diastolic dysfunction | 1 (10.0) | 0 ( 0.0) | 1 ( 5.3) |
| No left ventricular diastolic dysfunction | 7 (70.0) | 7 (77.8) | 14 (73.7) |
| Missing data | 2 (20.0) | 2 (22.2) | 4 (21.1) |
| Pericardial Effusion (n (%)) |  |  |  |
| Pericardial effusion | 2 (20.0) | 3 (33.3) | 5 (26.3) |
| No pericardial effusion | 6 (60.0) | 5 (55.6) | 11 (57.9) |
| Missing data | 2 (20.0) | 1 (11.1) | 3 (15.8) |

## DISCUSSION

In this cross-sectional study of 19 cases of suspected COVID-19 myocarditis, there was no prior history of coronary artery disease, atrial fibrillation, or heart failure; 21.1% of patients died; and 78.9% of cases required supplemental oxygen. In exploratory analysis comparing laboratory values at the time of diagnosis, a significantly higher geometric mean D-dimer and ferritin were observed in patients with fulminant compared to non-fulminant suspected myocarditis. On review of available echocardiography, 26.3% of cases had pericardial effusions. 10 out of the 16 with available echocardiographic data had normal left ventricular systolic function. The absence of gross differences in ejection fraction across fulminant and non-fulminant cases were difficult to interpret because of missing data in the setting of a small sample size.

The present study characterizes clinical COVID-19 myocarditis through descriptive analysis. Clinical COVID-19 myocarditis is an important entity to characterize because its signs and symptoms overlap with sepsis-induced cardiomyopathy, demand mediated ischemia, and ARDS but it may have unexplored and distinct clinical implications.^4^ As a relatively small cross-sectional study from a convenience sample, this study is limited to generating hypotheses; statistical inference to a generalized population cannot be made. Significance on hypothesis testing can be interpreted as a measure of statistical extremes that are unlikely to be due purely to chance, which merits further investigation and can motivate variable selection in future studies.

On average, troponins were elevated 101 times the normal limit at the time of diagnosis. In prior research, 17% of patients with COVID-19 had troponin elevations above the 99th percentile.^12^ Meanwhile, only 2.9% of patients hospitalized with influenza had troponin elevations.^11^ The proportion of patients with troponin elevation and the clinical severity of these elevations is notable. In all patients, acute coronary syndrome was not suspected by chart review. This strengthens the hypothesis that cardiac damage is uniquely associated with COVID-19. As the absolute values of troponin elevation are remarkable in this cross-sectional study, further research may investigate the clinical significance of different levels of troponin elevation.

The geometric mean but not the arithmetic mean of D-dimer and ferritin were significantly higher in patients with suspected fulminant compared to non-fulminant myocarditis. Direct associations are difficult to infer due to the lack of a randomly sampled population in this study. Still, this raises the hypothesis that fulminant myocarditis may be mediated through higher systemic inflammation. Cases of viral myocarditis have been diagnosed both with and without viral particles observed on endomyocardial biopsy.^15^ The observed dose response association between clinical severity of myocarditis and inflammatory markers on a multiplicative scale further motivates investigation into an indirect mechanism of cardiac injury mediated through systemic inflammation.

In summary, the present study better characterizes the entity of clinically suspected COVID-19 myocarditis and substantiates the importance of investigating the extent to which COVID-19 is mediated by direct viral injury and/or indirect systemic inflammation through exploratory analysis. Future studies may investigate whether direct versus indirect mechanisms predominate and how this difference impacts clinical outcomes.

## Data Availability

Data is available for review upon reasonable request.

